# Adipo-B Index as a Novel Predictor of Beta-Cell Secretory Function Without Measuring Insulin: Cross-Therapeutic Validation Against C-Peptide Index in Treatment-Naïve Type 2 Diabetes

**DOI:** 10.64898/2026.10.24.26366378

**Authors:** Eiji Kutoh, Alexandra N. Kuto

## Abstract

**OBJECTIVE:** The adipo-B index (adipo-IR divided by HOMA-B) was introduced as an integrator of adipose tissue insulin resistance and pancreatic beta-cell function. Algebraic simplification reveals that insulin cancels from the formula, yielding FFA × (FBG–63)/360 — a composite of circulating free fatty acids and ambient glycemia. We investigated whether this index predicts beta-cell secretory capacity measured by HOMA-B and C-peptide index (CPR-index) across four therapeutic strategies.

**RESEARCH DESIGN AND METHODS:** A total of 253 treatment-naïve subjects with T2DM received monotherapy with modified Japanese diet for diabetes (MJDD, n=61), canagliflozin (n=67), pioglitazone (n=62), or sitagliptin (n=63). Correlations between baseline adipo-B and baseline beta-cell indices, and between their longitudinal changes, were examined. This is a prospective, non-randomized observational study.

**RESULTS:** At baseline, adipo-B showed significant negative correlations with HOMA-B with all the subjects. At 3 months, adipo-B significantly decreased and HOMA-B significantly increased with all the therapeutic groups. CPR-index significantly improved with canagliflozin and sitagliptin. Pioglitazone showed a trend toward improvement while MJDD demonstrated no changes. Changes in adipo-B (Δadipo-B) significantly and negatively correlated with changes in both HOMA-B and CPR-index in MJDD, canagliflozin, and sitagliptin. With pioglitazone, no such correlations were observed — consistent with the previously reported bidirectional regulation of insulin secretion by pioglitazone depending on baseline insulin secretory status.

**CONCLUSIONS:** The adipo-B index predicted beta-cell secretory capacity — assessed independently by conventional beta-cell indices — without requiring insulin or C-peptide measurement. These findings suggest adipo-B may serve as a cost-effective surrogate for assessment of beta-cell function in routine clinical settings.

**ARTICLE HIGHLIGHTS:**

- **Why did we undertake this study?** Insulin cancels algebraically from the adipo-B formula, questioning its validity. We tested if adipo-B predicts beta-cell function using the insulin-independent C-peptide index.
- **What is the specific question we wanted to answer?** Does adipo-B predict beta-cell secretory capacity (via HOMA-B and C-peptide index) without requiring insulin or C-peptide measurements?
- **What did we find?** Cross-sectionally, baseline adipo-B negatively correlated with HOMA-B and CPR-index. Longitudinally, Δadipo-B negatively correlated with ΔHOMA-B and ΔCPR-index for MJDD, canagliflozin, and sitagliptin—but not pioglitazone, reflecting its bidirectional beta-cell regulation.
- **What are the implications?** Adipo-B predicts beta-cell capacity without insulin or C-peptide testing, offering a cost-effective surrogate for routine and resource-limited clinical settings.

## OBJECTIVE

Pancreatic beta-cell function is central to the pathophysiology and clinical management of T2DM (1). Its assessment conventionally requires measurement of insulin or C-peptide — tests that are expensive, not universally available, and subject to inter-assay variability. A practical, cost-effective surrogate for beta-cell function would therefore have significant clinical value, particularly in resource-limited settings.

We previously introduced the adipo-B index [adipo-IR / HOMA-B= FFA × insulin /insulin × 360/(FGB-63) ] as a composite metric of the adipose tissue–pancreatic beta-cell axis (2). Algebraic simplification reveals that insulin cancels from the formula, yielding FFA × (FBG−63)/360 — a product of circulating FFA and ambient glycemia that reflects glucolipotoxic stress on the beta-cell. While this cancellation precludes a direct mechanistic claim of beta-cell measurement, it raises a question: does adipo-B nevertheless predict beta-cell secretory capacity independently?

To address this, we examined correlations between adipo-B and beta-cell function — assessed by both HOMA-B and C-peptide index — at baseline (cross-sectionally) and longitudinally across four distinct therapeutic strategies in treatment-naïve subjects with T2DM.

## RESEARCH DESIGN AND METHODS

### Ethics and Cohort

The study adhered to the Declaration of Helsinki, was approved by Gyoda General and Kumagaya Surgery Hospital IRBs, and obtained electronic informed consent. This is an extension of study UMIN000006860.

### Subjects

Between March 2019 and August 2026, 253 treatment-naïve T2DM subjects meeting Japan Diabetes Society criteria were enrolled (2). Exclusion criteria: creatinine ≥1.5 mg/dL, AST/ALT ≥70 IU/L, heart disease, hypertension >160/100 mmHg, T1DM, and pregnancy. This non-randomized, observational study compared four monotherapies: MJDD (n=61), canagliflozin (n=67), pioglitazone (n=62), and sitagliptin (n=63). Daily female/male doses were 50/100 mg for canagliflozin, 15/30 mg for pioglitazone, and 25/50 mg for sitagliptin (2).

### Biochemical Measurements

Fasting blood samples (glucose, insulin, FFA, C-peptide) were collected before breakfast. Indices were calculated as previously described (2-5):

- adipo-IR = FFA × insulin
- HOMA-B = insulin × 360 / (FBG − 63)
- adipo-B = adipo-IR / HOMA-B = FFA × (FBG − 63) / 360
- CPR-index = fasting C-peptide (ng/mL) × 100 / FBG (mg/dL)

### Statistical Analysis

Changes represent the difference between baseline and 3 months. Within-group changes were assessed via paired t-tests, and associations via simple linear regression using PAST version 5.3 (University of Oslo). Statistical significance was set at p<0.05; p-values between 0.05 and 0.10 indicated a trend toward significance (6).

## RESULTS

### Characteristics of subjects and changes in metabolic parameters

A total of 253 treatment-naïve subjects with T2DM were included, comprising 61 treated with MJDD, 67 with canagliflozin, 62 with pioglitazone, and 63 with sitagliptin. Baseline metabolic characteristics and 3-month values are shown in Table 1. Adipo-B significantly decreased and HOMA-B significantly increased across all therapeutic groups. CPR-index significantly improved with canagliflozin and sitagliptin. Pioglitazone showed a trend toward improvement (p=0.0501) while MJDD did not.

**Table 1.** Changes in metabolic parameters after 3 months of treatment with MJDD, canagliflozin, pioglitazone, and sitagliptin. Paired *t*-tests were used to compare baseline and 3-month values for each parameter. Values are presented as mean ± SD. A) MJDD; B) Canagliflozin; C) Pioglitazone; D) Sitagliptin.

| Treatment | Parameter | Baseline | 3 months | p-value |
| --- | --- | --- | --- | --- |
| A. MJDD | adipo-B | 0.246±0.154 | 0.203±0.145 | <0.02 |
|  | HOMA-B | 25.75±18.88 | 29.35±24.01 | <0.05 |
|  | CPR-index | 1.188±0.535 | 1.188±0.539 | n.s. |
|  | C-peptide (ng/mL) | 2.08±0.91 | 1.91±0.67 | <0.007 |
| B. Canagliflozin | adipo-B | 0.299±0.169 | 0.195±0.108 | <0.00001 |
|  | HOMA-B | 24.69±24.63 | 31.48±27.2 | <0.0002 |
|  | CPR-index | 1.084±0.630 | 1.271±0.665 | <0.0003 |
|  | C-peptide (ng/mL) | 1.93±1.01 | 1.79±0.92 | <0.04 |
| C. Pioglitazone | adipo-B | 0.332±0.189 | 0.208±0.146 | <0.00001 |
|  | HOMA-B | 18.99±26.68 | 26.85±36.81 | <0.03 |
|  | CPR-index | 0.987±0.879 | 1.149±0.921 | 0.0509 |
|  | C-peptide (ng/mL) | 2.02±1.33 | 1.90±1.18 | n.s. |
| D. Sitagliptin | adipo-B | 0.381±0.256 | 0.290±0.257 | <0.0002 |
|  | HOMA-B | 18.75±13.01 | 26.47±19.47 | <0.0002 |
|  | CPR-index | 0.9444±0.480 | 1.200±0.653 | <0.00001 |
|  | C-peptide (ng/mL) | 1.91±0.89 | 2.07±1.11 | <0.02 |

Values are presented as mean ± SD. n.s., not significant.

### Adipo-B and beta-cell function

At baseline, adipo-B showed significant negative correlations with both HOMA-B (R=−0.376, p<0.00001) and CPR-index (R=−0.313, p<0.00001) for all 253 subjects combined (Figure 1A and 1B). After 3 months (Table 2), changes in adipo-B (Δadipo-B) showed significant negative correlations with both ΔHOMA-B and ΔCPR-index in the MJDD, canagliflozin, and sitagliptin groups. No significant correlations were observed with pioglitazone.

**Table 2.** Correlations of changes in adipo-B with beta-cell function indices.

| **$\Delta$ adipo-B vs. $\Delta$ HOMA-B** | **R** | **p-value** |
| --- | --- | --- |
| MJDD | -0.271 | <0.04 |
| Canagliflozin | -0.592 | <0.00001 |
| Pioglitazone | -0.185 | n.s. |
| Sitagliptin | -0.491 | <0.00005 |

| **$\Delta$ adipo-B vs. $\Delta$ C-peptide index** | **R** | **p-value** |
| --- | --- | --- |
| MJDD | -0.385 | <0.03 |
| Canagliflozin | -0.661 | <0.00001 |
| Pioglitazone | 0.018 | n.s. |
| Sitagliptin | -0.631 | <0.00001 |

n.s., not significant.

**Figure 1.**
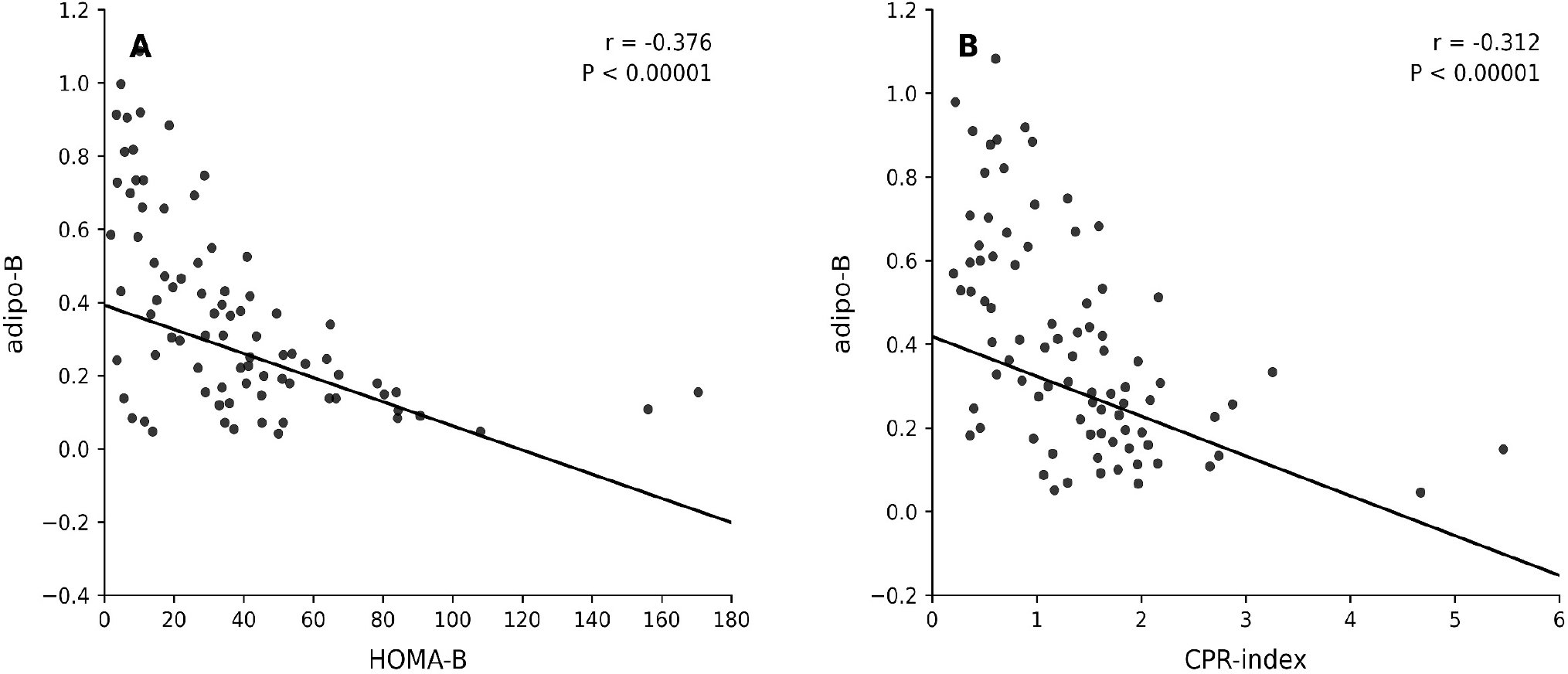
Correlations between changes (Δ) in adipo-B and changes (Δ) in beta-cell indices after 3 months of treatment. Simple linear regression analyses were performed between Δadipo-B and ΔHOMA-B or ΔCPR-index in each therapeutic arm: A) MJDD; B) canagliflozin; C) pioglitazone; D) sitagliptin.

## DISCUSSION

The present study demonstrates that the adipo-B index — despite the algebraic cancellation of insulin from its formula — retains meaningful biological validity as a predictor of beta-cell secretory capacity. At baseline, adipo-B showed highly significant negative correlations with both HOMA-B (R=−0.376) and CPR-index (R=−0.313) across all 253 subjects combined (Figure 1A and 1B), establishing it as a cross-sectional surrogate of beta-cell function without requiring insulin or C-peptide measurement. Longitudinally, Δadipo-B significantly and negatively correlated with both ΔHOMA-B and ΔCPR-index in three of four therapeutic arms (Table 2), demonstrating that adipo-B captures dynamic beta-cell recovery during treatment.

The biological rationale lies in the glucolipotoxicity framework. The simplified adipo-B formula — FFA × (FBG−63) / 360 — represents the combined burden of circulating FFA and ambient glycemia on the beta-cell. Chronic elevation of both FFA and glucose is well established to impair beta-cell function and survival through lipotoxic and glucotoxic mechanisms (7, 8). The strong negative correlation between adipo-B and CPR-index — a completely insulin-independent beta-cell measure — provides empirical validation of this glucolipotoxic interpretation, independent of the mathematical derivation of the index.

Crucially, the clinical utility of adipo-B relies on its retention of the glycemic component of HOMA-B, where (FBG − 63) represents the glucose excess above the theoretical beta-cell stimulation threshold (3). This specific mathematical feature distinguishes adipo-B from the triglyceride-glucose (TyG) index, which similarly correlates negatively with beta-cell function (9), in two important respects: adipo-B substitutes fasting FFA—the direct and proximal mediator of lipotoxicity—for triglycerides, and incorporates this biologically anchored glycemic offset rather than raw fasting glucose. Together, these elements offer a more mechanistically direct assessment of the glucolipotoxic burden on the beta-cell.

The strongest longitudinal correlations were observed with canagliflozin (Δadipo-B vs ΔCPR-index: R=−0.661, p<0.00001) and sitagliptin (R=−0.631, p<0.00001). Canagliflozin reduces adipo-B primarily through glucosuria-driven FBG reduction, simultaneously allowing beta-cell rest and functional recovery through a pathway independent of adipose tissue insulin resistance (2). Sitagliptin directly enhances incretin-mediated beta-cell function while reducing FBG and FFA (10), producing tightly coupled improvements in both adipo-B and CPR-index. With MJDD, caloric restriction reduces both FFA and FBG simultaneously, relieving glucolipotoxic stress and allowing beta-cell recovery in a physiologically coherent manner (2).

Notably, although the mean CPR-index did not change significantly with MJDD (Table 1), individual changes in adipo-B were significantly and negatively correlated with individual changes in the CPR-index (R = −0.385, p< 0.03, Table 2). This apparent discrepancy reflects heterogeneous beta-cell responses to dietary intervention: subjects with the greatest reductions in adipo-B tended to show relative improvements in the CPR-index, whereas those with minimal adipo-B reduction showed stable or declining values, resulting in no net group-level change. Crucially, Table 1 reveals a significant reduction in raw C-peptide levels within the MJDD group (p< 0.007). This stability of the mean CPR-index, despite falling raw C-peptide, is explained by the proportional reductions in both C-peptide secretion and fasting glucose during caloric restriction, which mathematically preserves the normalized index at the group level. This finding illustrates that adipo-B captures individual variations in beta-cell response to diet that are otherwise obscured by group mean analysis, highlighting its potential utility as a precision tool for monitoring therapeutic responses at the individual level.

The absence of significant correlation between Δadipo-B and ΔCPR-index with pioglitazone (Table 2) likely reflects the previously reported bidirectional regulation of beta-cell secretion by this agent — whereby patients with higher baseline insulin secretory capacity experience reductions while those with lower capacity experience restoration — resulting in opposing directional changes that mask any aggregate correlation (11). The unique decoupling of the adipo-B/beta-cell axis with pioglitazone might reflect its distinct PPAR-γ-mediated mechanism of beta-cell restoration (12), independent of glucolipotoxic stress reduction.

The principal clinical implication is that adipo-B may serve as a practical, cost-effective surrogate for beta-cell function assessment. Fasting FFA and glucose are inexpensive, universally available, and low-variability measurements compared with insulin or C-peptide assays — making adipo-B particularly applicable in clinical settings with limited laboratory infrastructure. The consistent longitudinal relationship between Δadipo-B and ΔCPR-index across three mechanistically distinct therapeutic classes (Table 2, Fig. 2) further suggests that adipo-B may be useful for monitoring beta-cell recovery during treatment, irrespective of the specific pharmacological mechanism employed.

**Figure 2.**
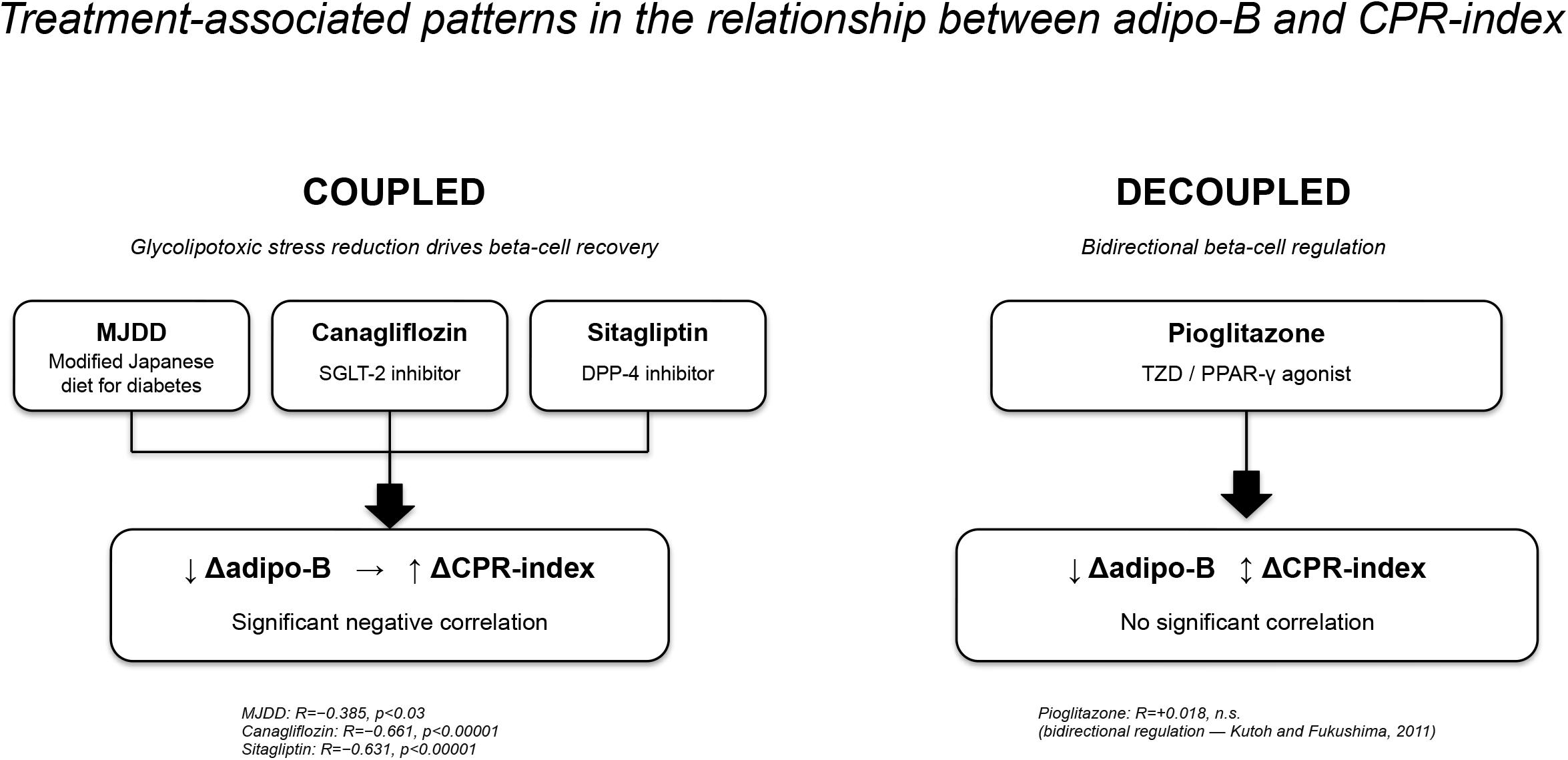
Schematic representation of the therapy-specific coupling and decoupling of the Δadipo-B/ΔCPR-index axis after 3 months of treatment.

Several limitations warrant consideration. This was a non-randomized observational study — treatment allocation was not controlled and confounding by indication cannot be excluded. Follow-up was limited to 3 months — long-term durability of the adipo-B/beta-cell relationship remains to be established. HOMA-B and CPR-index are validated surrogates but lack the precision of hyperinsulinemic-euglycemic clamp studies. The cohort comprised Japanese treatment-naïve subjects — generalizability to other ethnic populations requires independent validation. Larger, multi-center, randomized studies with longer follow-up are needed to confirm these findings and establish standardized clinical cutoff values for adipo-B.

## CONCLUSIONS

The adipo-B index predicts beta-cell secretory capacity — assessed independently by both HOMA-B and C-peptide index — without requiring insulin or C-peptide measurement. As adipo-B is derived solely from fasting FFA and glucose — inexpensive and universally available measurements — it may represent a cost-effective surrogate for beta-cell function assessment in routine clinical and resource-limited settings. The therapy-specific decoupling observed with pioglitazone further underscores the mechanistic complexity of beta-cell regulation across therapeutic classes and highlights the potential of adipo-B as a precision tool for individualized diabetes management.

## Data Availability

All data produced in the present study are available upon reasonable request to the authors

